# Cavernous Angioma Symptomatic Hemorrhage (CASH) Trial Readiness II: Imaging Biomarkers and Trial Modeling

**DOI:** 10.1101/2023.06.01.23290854

**Authors:** Stephanie Hage, Serena Kinkade, Romuald Girard, Kelly D. Flemming, Helen Kim, Michel T. Torbey, Judy Huang, John Huston, Yunhong Shu, Reed G. Selwyn, Blaine L. Hart, Marc C. Mabray, James Feghali, Haris I. Sair, Jared Narvid, Janine M. Lupo, Justine Lee, Agnieszka Stadnik, Roberto Alcazar, Robert Shenkar, Nicholas Hobson, Dorothy DeBiasse, Karen Lane, Nichole McBee, Kevin Treine, Noeleen Ostapkovich, Ying Wang, Richard E. Thompson, Carolina Mendoza-Puccini, James Koenig, Timothy Carroll, Daniel F. Hanley, Issam A. Awad

## Abstract

**Background:** Quantitative susceptibility mapping (QSM) and dynamic contrast enhanced quantitative perfusion (DCEQP) MRI sequences assessing iron deposition and vascular permeability were previously correlated with new hemorrhage in cavernous angiomas. We assessed their prospective changes in cavernous angiomas with symptomatic hemorrhage (CASH) in a multisite trial readiness project (clinicaltrials.gov NCT03652181).

**Methods:** Patients with CASH in the prior year, without prior or planned lesion resection or irradiation were enrolled. Mean QSM and DCEQP of CASH lesion were acquired at baseline, and at 1- and 2-year follow-ups. Sensitivity and specificity of biomarker changes were analyzed in relation to predefined lesional symptomatic hemorrhage (SH) or asymptomatic change (AC). Sample size calculations for hypothesized therapeutic effects were conducted.

**Results:** We logged 143 QSM and 130 DCEQP paired annual assessments. Annual QSM change was greater in cases with SH than in cases without SH (p= 0.019). Annual QSM increase by ≥ 6% occurred in 7 of 7 cases (100%) with recurrent SH and in 7 of 10 cases (70%) with AC during the same epoch, and 3.82 times more frequently than clinical events. DCEQP change had lower sensitivity for SH and AC than QSM change, and greater variance. A trial with smallest sample size would detect a 30% difference in QSM annual change in 34 or 42 subjects (one and two-tailed, respectively), power 0.8, alpha 0.05.

**Conclusions:** Assessment of QSM change is feasible and sensitive to recurrent bleeding in CASH. Evaluation of an intervention on QSM percent change may be used as a time-averaged difference between 2 arms using a repeated measures analysis. DCEQP change is associated with lesser sensitivity and higher variability than QSM. These results are the basis of an application for certification by the U.S. F.D.A. of QSM as a biomarker of drug effect in CASH.

## Introduction

Cerebral cavernous malformation, also known as cavernous angioma (CA) is a common hemorrhagic vascular anomaly of the human brain, affecting > 1 million Americans. Although the risk of first symptomatic hemorrhage from a CA is very low (<1% per year), cavernous angiomas with symptomatic hemorrhage (CASH) carry a ten-fold risk of rebleeding with serious clinical sequelae^1^. Asymptomatic bleeds in lesions can also herald future clinically relevant hemorrhage^2^. Several drugs have been shown in pre-clinical studies to decrease CA bleeding, and are poised for clinical testing^3^. Currently, clinical magnetic resonance imaging (MR) can detect a new bleed in CA, if performed within 4-6 weeks of hemorrhage but does not quantify the extent of bleeding. Awaiting new symptoms and performing MRI to detect a new bleed, can miss sub-clinical bleeds and the cumulative impact of repeated hemorrhages in lesions during follow-up. Clinical trials aimed at decreasing rates of recurrent symptoms cannot be realistically performed, with fewer than 200,000 CASH cases in the U.S. per year (meeting rare disease criteria), and low bleeding event rates requiring potentially thousands of patients to confirm a drug effect.

The assessment of novel therapies would greatly benefit from sensitive biomarkers that are biologically linked to bleeding in CAs, which would facilitate the assessment of drug effects on lesional hemorrhage. Vascular leak is a fundamental feature of CAs, mediating hemorrhage and the associated accumulation of non-heme iron in lesions. Pre-clinical studies demonstrated rescue of vascular permeability and decreased lesional iron deposition with ROCK inhibitors and statins^4–6^, B-cell depletion therapy^7^ and propranolol^8^ in murine CA models recapitulating the human disease. Hence these drugs would be expected to alter CA lesional iron deposition and permeability in vivo.

Quantitative susceptibility mapping (QSM) is a noninvasive MRI technique that assesses iron content by quantifying the magnetic susceptibility of local tissues. The QSM technique quantifies magnetic field changes caused by local susceptibility sources (such as iron) by performing a dipole field-to-susceptibility inversion from a single-orientation MRI acquisition after phase unwrapping and background field removal of phase images. QSM provides excellent depiction of brain lesions with iron deposition in patients with cerebral microbleeds, multiple sclerosis, brain tumors, intracranial calcifications and hemorrhages, and neurodegenerative diseases^9^. Despite the heterogeneity of lesional blood products at different stages of evolution, iron content in surgically excised CA lesions precisely reflected the measured iron concentration in the same specimens by mass spectrometry, and phantom studies have validated QSM assessment of ferric, ferrous and ferromagnetic iron concentrations^10^. Further, multisite validation of QSM acquisitions has been conducted using multiple instruments at three different institutions, using a QSM phantom with concentrations of gadolinium, corresponding to magnetic susceptibilities of 0, 0.1, 0.2, 0.4, and 0.8 ppm, confirming accuracy, precision and reproducibility^11^. Researchers in Chicago and New Mexico optimized a second technique, dynamic contrast enhanced quantitative perfusion (DCEQP) measure on MRI in human subjects, reflecting mechanistically postulated vascular hyper-permeability in CAs^12,13^.

Both QSM and DCEQP were implemented successfully in over 200 CA subjects and showed strong inter-observer agreement in QSM and DCEQP measurements, stability of both measurements in clinically stable lesions, and reproducibility across MRI instrument platforms^10,14,15^. As predicted by the conservation of mass hypothesis, CA lesions with greater permeability had higher lesional iron content (QSM)^16^, and lesional iron content was greater in older patients and in CAs with prior symptomtic hemorrhage (SH)^15^. More recent studies demonstrated a significant increase of mean lesional QSM and DCEQP in human CA lesions that had been stable clinically and on imaging for more than a year, and then manifested interval SH or growth during longitudinal follow-up, while these did not change in stable lesions^14^. There were tight, sensitive and specific thresholds of QSM and DCEQP increases in association with clinical events. Therefore, it is postulated that the QSM and DCEQP may be used as *in vivo* measures of hemorrhagic activity and vascular leak, respectively, and may reflect sensitive, clinically meaningful therapeutic effect in human CAs.

To date, only a small pilot study at a single site has assessed lesional QSM change during prospective follow-up of CA patients who had suffered a SH within the prior year^17^. The overall goal of the CASH Trial Readiness project (clinicaltrials.gov NCT03652181), was to define and characterize the population of CASH patients at multiple sites, who might participate in therapeutic clinical trials. In an accompanying paper^18^, we report prospective rates of clinical rebleeding and changes in functional status and quality of life assessments. Here, we report changes in lesional iron content and vascular permeability, assessed by QSM and DCEQP respectively, during longitudinal follow-up. We report the sensitivity and specificity of these biomarker changes to bleeding events in the same lesions, and examine various trial models based on postulated drug effects on clinical bleeding versus biomarker measures as surrogate outcomes.

## Patients and Methods

### Study Design, Subject Enrollment and Cases Contributing Paired Biomarker Data

The study protocol and baseline features of trial eligible CASH subjects were published previously^19,20^. Details of study design are presented in the **Supplemental Methods**. Eligible patients were 18 years or older, harboring one or multiple CA in their brain and had a SH within the prior year in a lesion without prior or planned treatment (resection or irradiation). Cases were excluded if they had a spinal lesion as source of SH, prior brain irradiation, contraindication for contrast agent administration or unwillingness to undergo research MRI studies, pregnancy in the past 6 months, breastfeeding, inability to verify SH with clinical and imaging review, homelessness and incarceration, or any reason for unlikeliness to return for follow-up visits^19^. Patients were removed from the study at completion of year 2 final follow-up, upon surgical resection of CASH lesion, upon last follow up or withdrawal date, or at end of study on November 1, 2022. Patients who suffered an adjudicated SH and failed to complete follow-up visit and research imaging studies at the end of the epoch were counted as an SH, with a fraction year of follow-up (up till the date of SH) for the purpose of compiling SH rates per year.

Enrolled subjects were followed for up to 2 years. Clinical and research MRI studies were also conducted at enrollment and at the end of each year of follow-up. An epoch of follow-up was defined as 1 year ±1 month, with the 1st epoch being from enrollment (baseline) to the year 1 follow-up, and the 2nd epoch from year 1 to year 2 follow-ups. At each visit (baseline, year 1 and year 2), the MRI was assessed for lesional changes in relation to clinical symptoms. Lesions were labeled as having a detected SH or asymptomatic change (AC) during the 1^st^ or/and 2^nd^ epoch. An SH was defined by new lesional hemorrhage documented by imaging and attributable to new or worsening symptoms. An AC was defined as lesional growth of ≥ 3mm or new subclinical bleeding on comparable T2 or T1-weighted images respectively^2,19^. The clinical and research MRI sequences^11,19^ were obtained in the same session. Any outside imaging obtained during the year for any reason was reviewed, to ascertain any lesional SH or AC during the prior year.

By November 1, 2022, 121 subjects were due to their Year 1 follow-up, and 102 completed it, including 95 who also underwent the research MRI studies at their visit (93.1 % of subjects contributed paired research imaging biomarker data in the 1^st^ epoch). Eighty subjects were due for their Year 2 follow-up, of whom 70 completed clinical follow-up and 59 completed the year 2 research MRI sequences (84.3% of subjects contributed paired research biomarker data in the 2^st^ epoch). In total, 277 QSM and 276 DCEQP sequences were acquired. Figure 1 shows the respective cohorts contributing to clinical and biomarker assessments, SH and AC in these cohorts, and SH and AC in the cases contributing paired QSM and DCEQP assessments. Respective contributions of research imaging at the multiple sites, and a comparison of cases with and without biomarker assessments are summarized in the Supplemental Tables 1-3.

**Figure 1.**
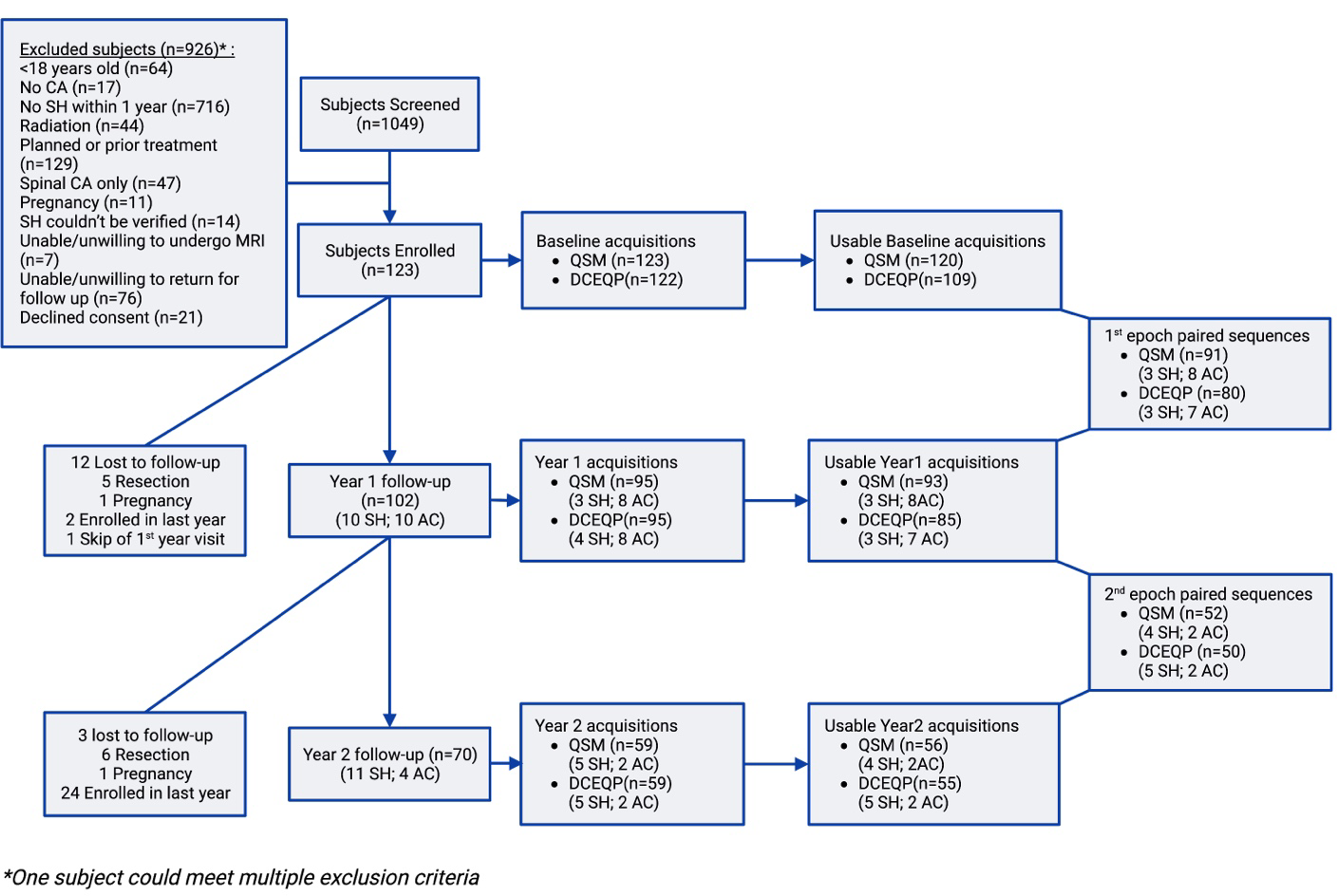
Flow chart of screened, excluded and enrolled subjects with yearly follow-up and contribution to research imaging acquisitions, usability and QSM/DCEQP pairs analyzed.

### Biomarker Data Acquisition and Post-processing

DCEQP and QSM scans across all five sites utilized a 3-Tesla field strength scanner with an eight-channel head coil for acquisition. The quantitative measurement of MRI biomarkers has been previously validated across instrument platforms and institutions using both static and dynamic phantom models^11^. Detailed protocols of research imaging have been previously published^11,14,19,21^ and are summarized in the **Supplemental Methods**. Figure 2 illustrates conventional MRI, QSM and DCEQP assessments of a CASH lesion. We excluded research sequences which were corrupted during acquisition, included artifacts, or did not cover the CASH lesion, and hence did not contribute usable QSM or DCEQP values.

**Figure 2.**
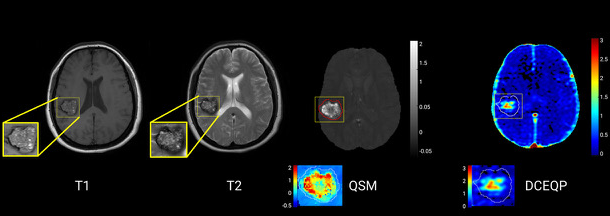
Illustration of conventional MRI axial images with (left to right) T1-weighted sequence, T2-weighted sequence, QSM, and DCEQP assessments of CASH lesion. The regions of interest for QSM and DCQEP assessments include the whole lesion as delineated on T2-weighted sequence. Note the maximal QSM values in the hemosiderin ring at the lesion periphery, while the maximal permeability values are within the core of the lesion.

### Definition of Biomarker Outcomes

Annual change in mean QSM and DCEQP of the CASH lesion were proposed as monitoring biomarkers of respective change in lesional iron content and permeability, to be targeted for potential treatment effect in clinical trials^17,19,22^.

Previous studies had established biologic plausibility and receiver operation curves (ROC) analyses of the QSM and DCEQP mean yearly change in initially stable CA (without a CASH event in the prior year), had identified an increase in mean lesional QSM of ≥ 5.81% (sensitivity 82.3%; specificity 88.89%) and an increase of ≥ 39.59% (sensitivity 78.72%; specificity 88.89%) in mean lesional DCEQP were associated with new SH during annual follow-up^14^. We thus defined and prespecified two categorical threshold “biomarker events” as ≥ 6% yearly change in mean lesional QSM and ≥ 40% annual change in mean lesional DCEQP.

### Statistical Analyses and Trial Simulations

Statistical analyses and sample size calculations in simulated clinical trials are detailed in the **Supplemental Methods**.

## Results

### Change in Mean Lesional QSM During Annual Follow-up of CASH Lesions

Raw data on mean lesional QSM assessments are presented in the **Supplemental Figure 1**. Of 295 potential QSM sequences due by the 1^st^ of November 2022 (123 at baseline, 102 at year 1, and 70 at year 2), 277 were conducted, hence 93.9% feasibility of QSM acquisition. Of 277 acquired sequences, 269 were usable (97.1%) and contributed to 143 total pairs of QSM (91 and 52 QSM pairs in the 1^st^ and 2^nd^ annual epochs of follow-up, respectively). There were 3 cases with SH and 8 with AC during the 1^st^ year, and 4 SH and 2 AC during the 2^nd^ year, among cases with corresponding paired QSM assessments (Figure 1). The overall mean QSM yearly-change was +17.52 (SD 73.27). QSM yearly-change during epoch 1 (mean +16.33; SD 74.93) was significantly lower than in epoch 2 (mean +19.61; SD 70.96) (p 0.033) with a correlation of ρ −0.39 (Figure 3). There were no significant differences in QSM yearly-change between cases with SH (mean +34.64; SD 27.17) and cases with AC (mean +38.53; SD 59.36). QSM changes in cases with SH, and those with SH or AC were significantly higher than QSM change in cases with no clinical events (p 0.019 and 0.0043 respectively) (Figure 4).

**Figure 3.**
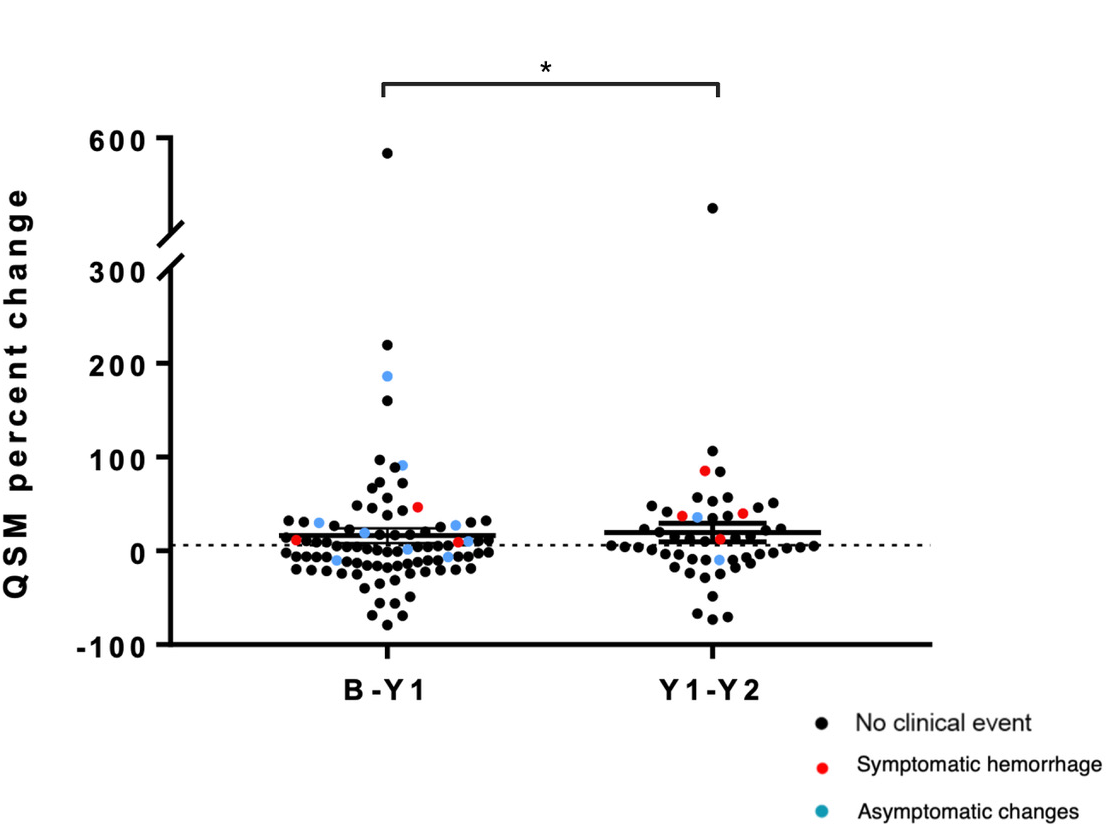
QSM percent change during the 1^st^ and 2^nd^ follow-up years. QSM yearly-change during epoch 1 (mean +16.33; SD 74.93) was significantly lower (p=0.033) than in epoch 2 (mean +19.61; SD 70.96). Red dots represent SH; blue dots represent AC; and black dots represent no clinical event detected.

**Figure 4.**
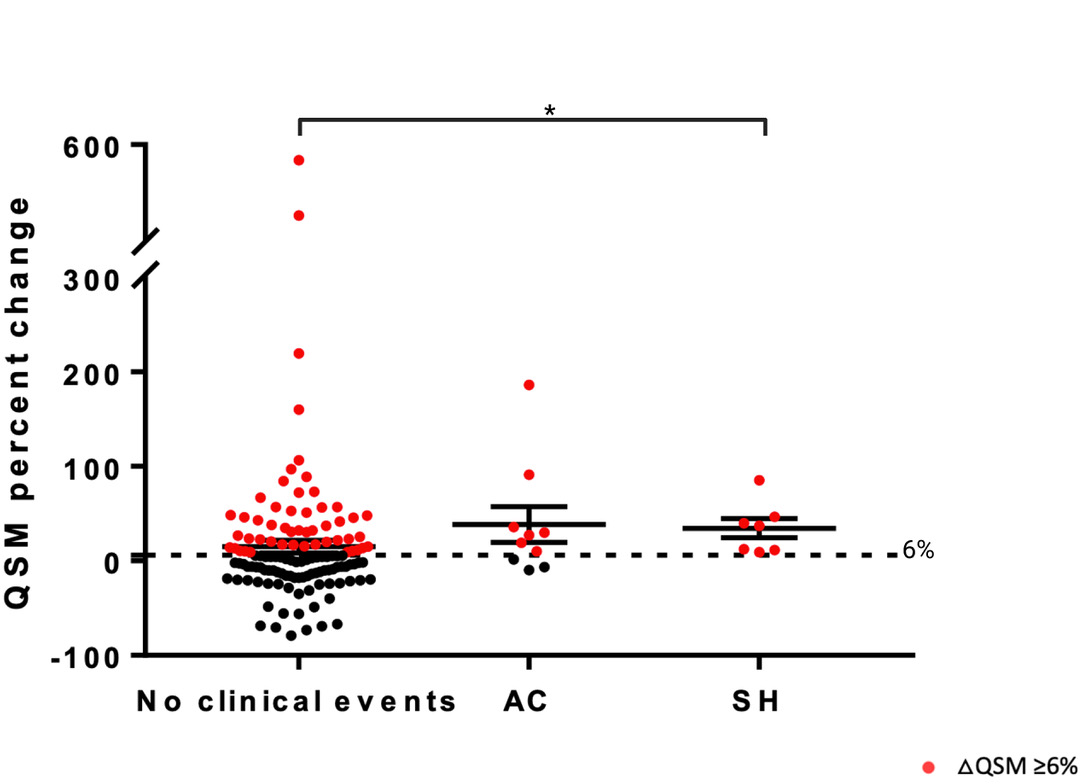
QSM yearly percent change for cases with SH, AC and no clinical events detected during the same epoch as paired biomarker assessment. No significant differences in percent yearly-change in mean lesional QSM between cases SH (mean +34.64; SD 27.17) and AC (mean +38.53; SD 59.36) during the same epoch. QSM changes with SH events were significantly higher than QSM change in cases with no clinical events (p 0.019). Red dots represent QSM pairs that meet the threshold biomarker ΔQSM≥6%.

A QSM increase by ≥ 6% was found in all 7 SH cases contributing paired QSM assessments, indicating 100% sensitivity for recurrent SH. A QSM increase by ≥ 6% occurred in 7 of 10 AC cases, with a sensitivity of 70% to ACs identified in the study. The 3 cases categorized as AC per prespecified criteria (increase of T1 signal or hemorrhagic lesion growth by > 3 mm) without QSM increase by ≥ 6% may have represented evolution of prior bleed rather than new bleeding, hence an imprecision of the definition of AC by predefined MRI criteria (**Supplemental Figure 2**). A QSM increase by ≥ 6% was documented in 51 pairs without SH or AC, a relative frequency 3.82 greater than the occurrence of clinical events.

### Change in Mean Lesional DCEQP During Annual Follow-up of CASH Lesions

Raw data on mean lesional DQEQP assessments are presented in the **Supplemental Figure 1**. Of 295 potential DCEQP sequences, 276 were acquired, hence a feasibility rate of 93.6%. Of these, 249 sequences were usable after processing, with a usability rate of 90.2%, contributing 130 DCEQP annual paired assessments. Three SH and 7 AC occurred during the 1^st^ epoch, and 5 SH and 2 AC during the 2^nd^ epoch among cases with corresponding paired DCEQP assessments (Figure 1). Mean lesional DCEQP yearly-change was +58.09 (SD +232.6). DCEQP yearly-change during the 1^st^ epoch (mean +61.62; SD +248.8) was not significantly different from the change during the 2^nd^ epoch (mean +52.45; SD +206.4) with a correlation of ρ −0.15 (Figure 5). DCEQP pairs with SH events had a higher mean lesional change (mean +179.9; SD +237.4) than DCEQP pairs without detected clinical events (mean +49.92; SD +235.5) (p 0.030). There was no significant difference in yearly DCEQP change in cases with AC events (mean +52.45; SD +175.8) and SH events (Figure 6). DCEQP annual increase was found in 6 of the 8 cases with recurrent SH cases indicating 75% sensitivity, and in 3 out of 9 AC cases with a sensitivity of 33.33%.The DCEQP increase ≥ 40% was identified in 43 pairs without SH or AC, 3 times more frequently than clinical events.

**Figure 5.**
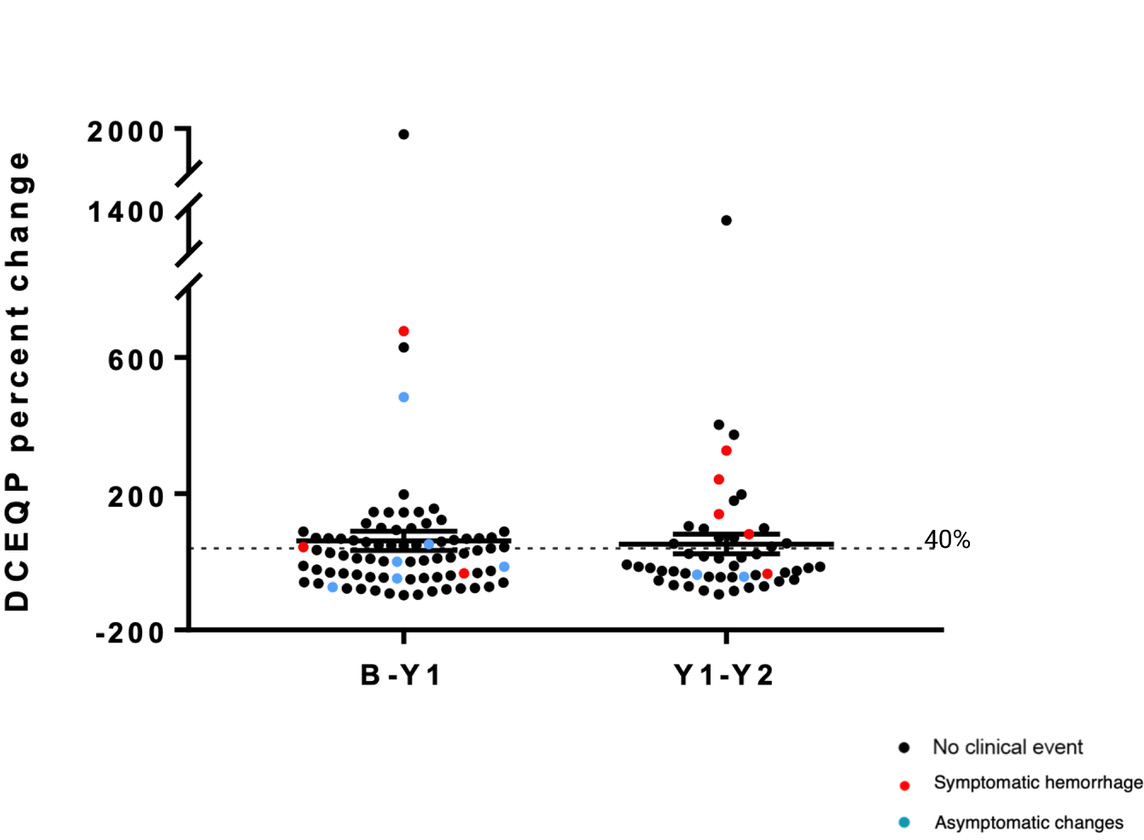
DCEQP percent change during the 1^st^ and 2^nd^ follow-up years. DCEQP yearly-change during the epoch 1 (mean +61.62; SD +248.8) was not significantly different from the change during epoch 2 (mean +52.45; SD +206.4). Red dots represent SH; blue dots represent AC; and black dots represent no clinical event detected.

**Figure 6.**
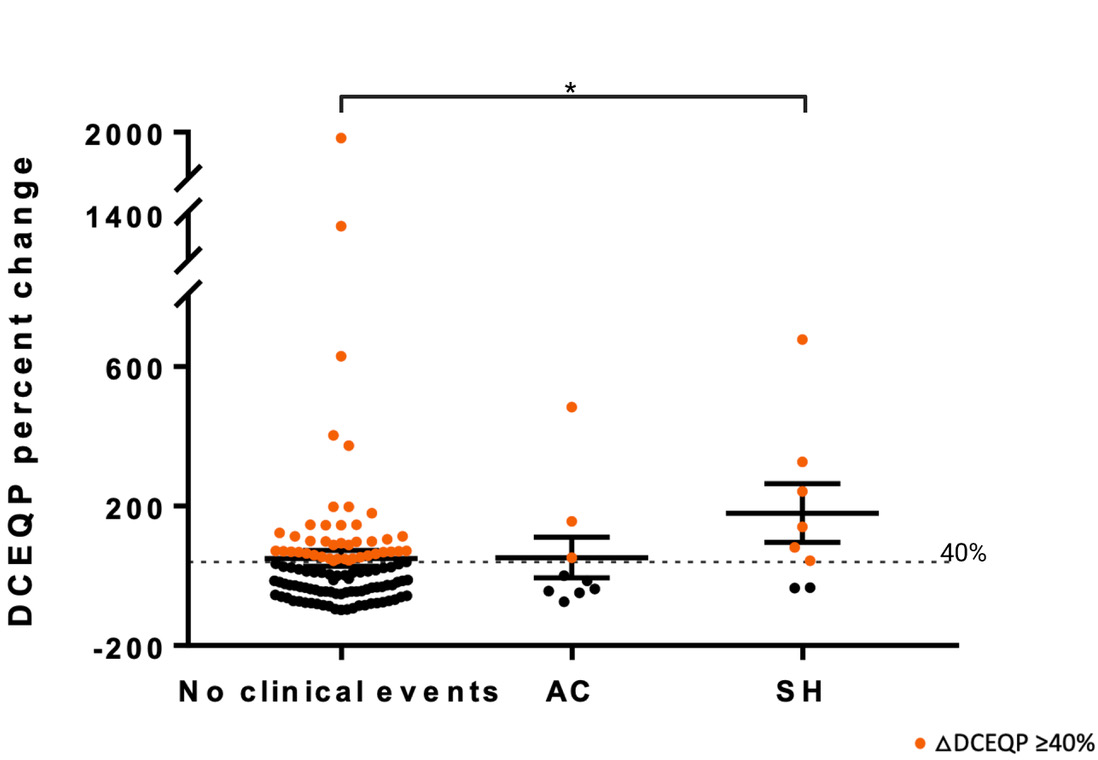
DCEQP yearly percent change for SH, AC and no clinical events detected during the same epoch as paired biomarker assessment. There was significantly higher percent change in mean lesional DCEQOP (mean +179.9; SD +237.4) in cases with SH events than in those without detected clinical events (mean +49.92; SD +235.5) (p 0.030). No significant difference in yearly DCEQP change between AC events (mean +52.45; SD +175.8) and either SH events or cases with no clinical events. Orange dots represent DCEQP pairs that meet the threshold biomarker ΔQSM≥40%.

### Trial Modeling

Sample sizes for potential trials were based on variance and co-correlation of % biomarker changes in 1^st^ and 2^nd^ epochs, a hypothesized therapeutic effect on mean lesional QSM or DCEQP change, a time-averaged difference between the two groups using a repeated measures analysis conducted as an unadjusted linear mixed model^17,19,22^, assumed power of 0.8 and alpha of 0.05 (two-tailed or one-tailed), 1:1 randomization, and patient follow-up for two years (2 paired biomarker assessments). For an interventional effect of 30% reduction in QSM change, a trial population of 42 (two-tailed) or 34 (one-tailed) subjects are sufficient, respectively (Supplemental Table 4) with two years follow-ups and two paired biomarker assessments per subject. Detecting a 20% reduction in QSM change would require 92 subjects if two-tailed test, and 72 subjects if one-tailed test. Because the variances and consequently the standard deviations of DCEQP yearly-change in both epochs were considerably higher, a clinical trial using DCEQP as a surrogate outcome would require hundreds to thousands of subjects in order to demonstrate a therapeutic effect (Supplemental Table 4).

We also considered a composite categorical outcome of any SH, AC, or QSM biomarker increase ≥ 6%. This would effectively count cases with SH or AC who did not undergo biomarker assessments in the epoch where they suffered the SH or AC, as well as those whose lesion showed a QSM increase ≥ 6%, (shown to be sensitive to lesional bleeding and more frequent than SH and AC events). There were overall 21 cases with SH in the trial readiness follow-up and biomarker validation cohort^18^, and 86 with SH, AC or QSM ≥ 6% during 175 patients-years of follow-up. The annual SH rate in this multisite clinical trial was 12.0%, and this composite outcome rate was much more frequent at 49.2%. Trial sample sizes based on a change of SH rate alone versus composite outcome rate (SH, AC or QSM ≥ 6%) are presented in the Supplemental Table 5. To demonstrate a 30% reduction in SH rates, total sample sizes of 2218 (two-tailed) or 1746 (one-tailed) patient-years of follow-up are required, assuming a power of 0.8 and alpha of 0.05 (two-tailed or one-tailed), and 1:1 randomization. A 30% reduction of the composite outcome rate can be detected with 356 (two-tailed) or 274 (one-tailed) patient-years, or 178 and 137 patients followed for two years, assuming the same event rates in both years.

## Discussion

We here demonstrate the feasibility and usability of QSM and DCEQP acquisitions during prospective follow-up of trial eligible CASH subjects. Among patients who came to the research sites for scheduled visits, 93.9% and 93.6% completed QSM and DCEQP acquisitions, respectively. Of these, 97.1% and 90.2% were usable. This is very encouraging regarding potential imaging biomarker implementation in clinical trials. This likely reflected protocolized site initiation and education, phantom assessment of QSM and dynamic T1-weighted signal at each MRI instrument, the chaperoning of research imaging acquisition, including repeat acquisitions in cases of head movement or other technical problems^11^. All imaging research sequences were post-processed at the Chicago imaging core. It is unclear if similar biomarker performance could be achieved without these rigorous protocols.

Published correlations of QSM and DCEQP changes in CAs had mostly addressed previously stable lesions^14^. Changes in CASH lesions would be expected to be more complex, as they include recovery from recent hemorrhage and greater likelihood of rebleeding. To date, only a small pilot study had assessed lesional QSM change during prospective follow-up of patients who had suffered a SH within the prior year^17^. During 22 patient-years of follow-up in 16 subjects, there were twice as many cases with a documented threshold biomarker event (≥ 6% QSM increase) as clinical events, and no clinical event occurred without such threshold increase in QSM. We here confirmed greater increases in QSM and DCEQP changes in lesions with a repeat SH or AC during the annual follow-up than in those without clinical events. We also confirmed an annual QSM change ≥ 6%, which had been associated with new SH in previously stable CAs, in every case with new SH during the same year (100% sensitivity). Sensitivity was lower for AC in lesions, although this partly suggests imprecise definition of new bleeding versus evolution of prior bleeding based on conventional MRI features. Lower sensitivity was observed with DCEQP changes, and far greater variances.

An effect of atorvastatin versus placebo on percent change in mean lesional QSM in CASH lesions has been postulated as a primary outcome of the ongoing prospective double-blind Phase I-IIA proof of concept trial (clinicaltrials.gov: NCT02603328), where patient enrollment has been completed and follow-up is underway of enrolled cases for two years^22^. A prior pilot study with a small number of cases addressed the effect of simvastatin on DCEQP in CA lesions^23^. However, that study was underpowered and did not address CASH lesions, nor drug equivalent doses required for effect in preclinical studies^5,6^.

Based on results herein at multiple sites, trial simulations endorse a very efficient sample size by using mean lesional annual percent QSM change as an outcome, and a time-averaged difference between the two groups using a repeated measures analysis, conducted as an unadjusted linear mixed model^17,19,22^. With this approach, the most efficient trial model would detect a 30% relative risk reduction in QSM annual change with only 34 or 42 subjects (one and two-tailed, respectively), followed for two years, 1-1 assignment, power=0.8, and alpha=0.05. This is clinically meaningful and in line with effect sizes on iron deposition in preclinical models ^4–8^. Such efficient sample sizes are attractive for platform trials, and for assessing the comparative effects of multiple drugs or doses in a rare disease. Two-tailed trials would be needed for potential drugs where there may be concern about increasing or decreasing lesional bleeding (i.e., statins, aspirin), while one-tailed trials are applicable for drugs where there is no concern about increasing hemorrhage (i.e. propranolol). The results herein motivated an application under review by the United States Food and Drug Administration, Center for Drug Evaluation and Research Biomarker Qualification Program Drug Development Tool (DDT-BMQ-000127) to assess drug effects on lesional bleeding in CAs.

Lower feasibility, usability and sensitivity for SH, very high variances, and the need for repeated gadolinium administration, dampen enthusiasm for using DCEQP as a similar biomarker. In fact, sample size calculations endorsed the need for huge sample sizes, in comparison to QSM when targeting an effect on DCEQP change.

Since these simulations are derived from cases with paired QSM or DCEQP assessments, there is concern that they would not reflect cases with missing paired biomarker assessment, particularly cases with known SH or AC who underwent surgery or otherwise did not complete post-bleed biomarker assessments. Indeed, cases with SH were less likely to contribute paired biomarker assessments (Supplemental Table 1), possibly due to undergoing surgery after a documented SH. Also, males were less likely to contribute paired QSM assessments. One possible approach to adjust for missing biomarker data would be to impute the mean QSM change of cases with no clinical events in cases missing biomarker data and who had no SH/AC events, and the mean QSM change of SH or AC in cases missing biomarker data who had SH or AC events. Another option is to consider a composite outcome of any SH or AC or QSM biomarker increase ≥ 6%. This would effectively count cases with SH or AC who did not undergo biomarker assessments in the epoch where they suffered the SH or AC, as well as those whose lesion showed a QSM increase ≥ 6%, shown to be sensitive to lesional bleeding and biologically plausible. SH annual rate in this multisite clinical trial was 12.0% and this composite annual outcome rate was much more efficient 49.2%. A 30% relative risk reduction in two groups on the composite annual outcome rate can be detected with 356 (two-tailed) or 274 (one-tailed) patient-years, more efficient than with SH alone, but clearly less efficient than modeling on % lesional QSM change.

We only assessed changes in QSM and DCEQP during annual epochs of follow-up, and we cannot comment about the value of more frequent assessments. It is also unclear what is the specific timeline of increase of QSM or DCEQP in lesions after a bleed, or its recovery. Among cases who completed paired biomarker assessments, SHs tended to cluster toward the end of the epoch (Supplemental Figure 3). This could be explained by greater likelihood of SHs undergoing surgery earlier in the epoch, and not completing paired biomarker assessments (Supplemental Table 1). Despite this clustering, there was a trend toward greater percent mean lesional QSM and DCEQP change when the SH occurred closer to the to the second biomarker acquisition, consistent with greater sensitivity to more recent bleeding.

We prespecified a percent change in annual mean lesional biomarker measures, based on pilot studies where absolute changes, median, maximum or minimum values were not as sensitive or specific in relation to clinical bleeding^14,15^. The lesional border as defined on T2-weighted sequences was used for the assessments of mean values of imaging pixels comprising the lesional region of interest. We often that maximal QSM areas often involved the CA periphery, and DCEQP maximal values often involved the core of the lesion (Figure 2), although there were substantial heterogeneities (**Supplemental Figure 2**). More recently, Incerti, et al. reported maximal lesional QSM as a prognostic biomarker of bleeding in CAs,^24^ but they did not specify how many pixels were considered for such maximal values, nor did they analyze mean lesional QSM or report changes in QSM over time. Sone, et al. from our team analyzed multiple derivations of permeability and perfusion (mean, median, upper or lower terciles, coefficient of variation, skewness, kurtosis, entropy, high-value cluster mean and area, as well as low-value cluster mean and area) from DCEQP acquisitions at a single point in time, as diagnostic and prognostic biomarkers of CASH^25,26^. In ongoing studies outside the scope of this report, we propose to analyze changes of these other biomarker derivations over time in the Trial Readiness cohort, as well as exploring artificial intelligence analysis of other features of QSM and DCEQP maps, and their correlations with changes in plasma biomarkers.

## Data Availability

All data referred to in the manuscript is available through the corresponding author.

## Non-standard Abbreviations and Acronyms<colcnt=2>

AC: asymptomatic change
CA: cavernous angioma
CASH: cavernous angiomas with symptomatic hemorrhage
DCEQP: dynamic contrast enhanced quantitative perfusion
MRI: magnetic resonance imaging
QSM: quantitative susceptibility mapping
SH: symptomatic hemorrhage
SD: standard deviation

## Acknowledgments

None

## Supplemental Materials

Supplemental Methods

Supplemental Figures and Figures Legends

Supplemental Tables

References 27-28

## Supplemental Methods

### Follow-up and biomarkers validation cohort

This is an observational multisite cohort study with enrollments of trial eligible CASH patients starting on 8/15/2018 through 12/17/2021, with planned annual follow-ups for up to 2 years. The study was funded by the National Institute of Neurological Disorders and Stroke (UO1NS104157; Clinicaltrials.gov NCT03652181), and its protocol was published previously.^19^ Screening and enrollment rates, and baseline features of trial eligible CASH patients were previously published.^20^ The study was overseen by a Central Institutional Review Board at the University of Chicago Medicine. Data monitoring and quality reviews were performed by research staff at Johns Hopkins University.

The follow-up and biomarker validation (FUBV) cohort reported herein was enrolled at 5 high volume CA programs at the University of Chicago Medicine, Mayo Clinic Rochester, University of California San Francisco, University of New Mexico and Johns Hopkins Medical Center. Sample size and power calculations were generated based on NQuery Advisor version 7. Assuming a sample size of 120 CASH cases, and allowing 30% dropouts or missing major biomarker or other data, we would have 84 evaluable patients (168 patient-years).^19^

Initially planned to follow 120 subjects for two years, the target follow-up was changed by the Executive Committee to include all cases who completed at least one follow-up by November 1, 2022, in view of delays during the Covid-19 pandemic and the end of project funding. During the Covid-19 pandemic, some clinical follow-ups were completed virtually, hence these patients did not contribute to research biomarker imaging. In the end, the FUBV cohort included 121 subjects who completed Year 1 clinical follow-up and 70 who completed year 2 clinical follow-ups (191 patient-years), and 154 completed paired annual biomarker assessments.

### Research sequences, imaging biomarker acquisitions at multiple sites, and postprocessing

Respectively, 75.5 %, 12.6 %, 7.0%, 2.8 %, and 2.1% of the research magnetic resonance imaging (MRI) sequences were acquired at the University of Chicago, Mayo Clinic Rochester, University of New Mexico, University of California San Francisco, and Johns Hopkins University (the latter two sites were activated later in the study, to enhance recruitment during the pandemic). Demographics or clinical features cases with and without contribution of at least one paired biomarker imaging were not different at the respective sites, except for a greater prevalence of recurrent symptomatic hemorrhage (SH) cases (p=0.027) and males (p=0.037) among patients failing to contribute paired research imaging sequences (Supplemental Results).

The MRI models used included the Achieva and Ingenia Elition 3.0T X (Philips Medical Systems, Best, The Netherlands), as well as the MAGNETOM Skyra and Prisma (Siemens Healthcare, Munich, Germany). The quantitative susceptibility mapping (QSM) and dynamic contrast enhanced quantitative perfusion (DCEQP) data sets were acquired by trained imaging scientists at each site as per published protocols I^11^, and the deidentified data was securely shared using a secure web-based electronic data capture system, VISION (Prelude Dynamics, Austin, Texas) hosted by Johns Hopkin’s University’s BIOS Clinical Trial Coordinating Center. All clinical and research MR sequences were anonymized by the site where imaging was completed to protect patient health information. The anonymized sequences were then uploaded to a secure Electronic Data Capture (EDC) system (VISION) hosted by Johns Hopkin’s University’s BIOS Clinical Trial Coordinating Center.

Imaging data were transferred to the University of Chicago where post-processing and QSM and DCEQP data was generated. QSM acquisitions were generated using a single 3D, 8 echo, spoiled gradient recalled echo (GRE) T2*-weighted sequence (acquisition time 11 minutes), using a previously published detailed protocol^14,15^. Vascular permeability was measured via a DCEQP protocol utilizing seven pre-contrast T1-weighted sequences with varying time delay (TD) values from 120 ms to 10 s, followed by a gadolinium-enhanced (MultiHance; gadobenate dimeglumine, Bracco Diagnostics, Inc.) dynamic scan of 250 frames with a time resolution of 1.2 s (acquisition time 9 minutes including set-up). 5 axial T2-weighted turbo spin echo (TSE) images of high spatial resolution were captured to serve as an anatomical reference when selecting lesion regions if interest (ROIs).

These data were processed using MATLAB software (MathWorks, Natick, MA) and the operators were blinded to the clinical status of patients throughout image analysis. Quantitative iron susceptibility was calculated using a Morphology Enable Dipole Inversion (MEDI) Toolbox (Medimagemmetric, LLC, New York City, New York) to generate a QSM map^27,28^. Mean lesional QSM values were generated by selecting ROIs around the lesion on all QSM map slices where the lesion is visible/present, shape and size confirmed by referencing a T2-weighted sequence, then averaging all the voxels inside the ROI^11^.Vessel permeability was calculated using a previously published protocol utilizing Patlak’s voxel-wise method and a two-compartment model to generate a blood brain barrier permeability map^21^. Mean lesional DCEQP values were generated by selecting an ROI around the lesion on the Ax T2 TSE slice that contained the largest lesion module and hemosiderin ring, then averaging those voxel values within the blood brain barrier (BBB) permeability map. Post-processing was completed by clinical research fellows and research technicians at the University of Chicago, following an established protocol with high intra- and inter-observer agreement^13,15^. All ROIs were adjudicated by a clinical research fellow (S.H.) and were selected using previously published protocol^10,11^.

### Statistical analyses, sensitivity and relative frequency

Mean lesional changes in QSM and DCEQP were calculated by relative percent changes for each annual epoch as continuous variables:

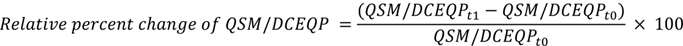

t0: 1^ST^ point in the epoch

t1: 2^nd^ point in the epoch

Cases with pairs of imaging biomarkers at the beginning and end of the annual follow-up epoch were included. Analyses were applied on annual percent change in lesional QSM and DCEQP. Paired Wilcoxon t-test compared the difference of annual change of mean lesional QSM and permeability during the 1^st^ and the 2^nd^ annual epochs. Mann Whitney t-test compared the annual change of mean lesional QSM and permeability in lesions with no clinical events and lesions with a clinical event (SH or asymptomatic change [AC]) documented during the corresponding epoch-year. The sensitivity and relative frequency of the threshold biomarker events were assessed based on their rate of occurrence and the rates of SH and AC during the same epochs with paired biomarker acquisitions.

A highly sensitive biomarker event means few false negative results, thus few clinical events would be missed by monitoring biomarker change and this is now confirmed for QSM change ≥ 6% in relation to SH.

Our goal was the detection and potential therapeutic targeting of occult and symptomatic bleeding in CAs. Hence, we did not define the absence of SH or AC as true negative events, and we did not aim to assess the specificity of biomarker changes, or their ability to designate an individual who does not have an event as negative.

### Trial simulation and sample size calculations

Based on variance and co-correlation of % biomarker changes in 1^st^ and 2^nd^ epochs, sample size calculations were conducted to power trials for a hypothesized therapeutic effect on mean lesional QSM change or mean lesional DCEQP change. We hypothesized a time-averaged difference between the two groups using a repeated measures analysis that was conducted as an unadjusted linear mixed model^17,19,22^. The effect sizes modeled were based on the minimum clinically significant changes between groups (relative risk reduction of 20%, 25% and 30%), as documented in preclinical studies ^4,5,22^. We used the within patient-correlation and the standard deviation (SD) of the mean percent change for QSM and DECQP for each epoch observed in our study to estimate the SD of the mean change in each measure over both epochs. In estimating the sample sizes, we assumed power of 0.8 and alpha of 0.05 (two-tailed or one-tailed), 1:1 randomization, and patient follow-up for two years (2 paired biomarker assessments). The power of future trials based on hypothesized intervention effect on the annual rate of SH or on a composite outcome (SH, AC or QSM threshold biomarker event) was based on the two sample test of proportions, and also assumed a power of 0.8 and both the two-tailed and one-tailed alpha of 0.05.

### Potential confounders

Supplemental Table 1 shows fewer cases with research sequences among males and those with SH. Younger patients contributed paired biomarker assessments at University of Chicago than other sites. The newest and lowest enrolling site contributed significantly less year 2 paired assessments than other sites.

Cases with SH who underwent paired biomarker assessments tended to have their bleeds later in the epoch (Supplemental Figure 3). Despite this clustering and the small number of cases with SH at various time points within the epoch, there was a trend toward greater percent mean lesional QSM and DCEQP change when the SH occurred closer to the to the second biomarker acquisition.

## Supplemental Figures and Figures Legends

### Mean lesional QSM and DCEQP values at each point in time

**Supplemental Figure 1A:**
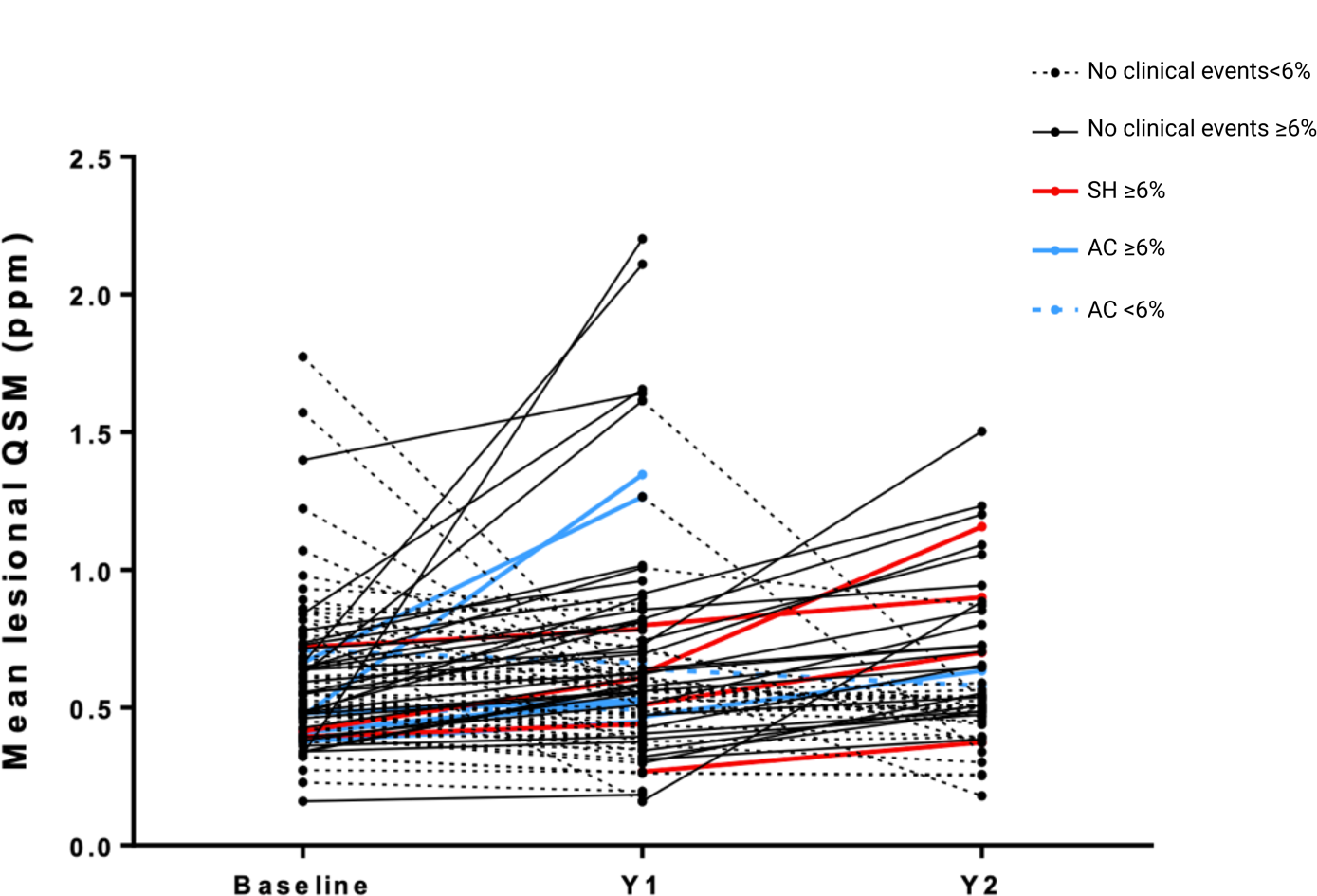
Mean lesional QSM at baseline and at 1st and 2nd follow-up years. Black lines represent QSM pairs with no clinical events. Red lines represent QSM pairs with an SH event. Blue lines represent QSM pairs with AC events. QSM pairs meeting the threshold biomarker are solid lines. QSM pairs not meeting the biomarker threshold are dashed lines. Red lines represent yearly-epochs with SH; blue lines represent AC; solid lines represent epoch-years that met the threshold of ΔQSM≥6% and dashed lines are for the ones that did not meet the threshold biomarker.

**Supplemental Figure 1B:**
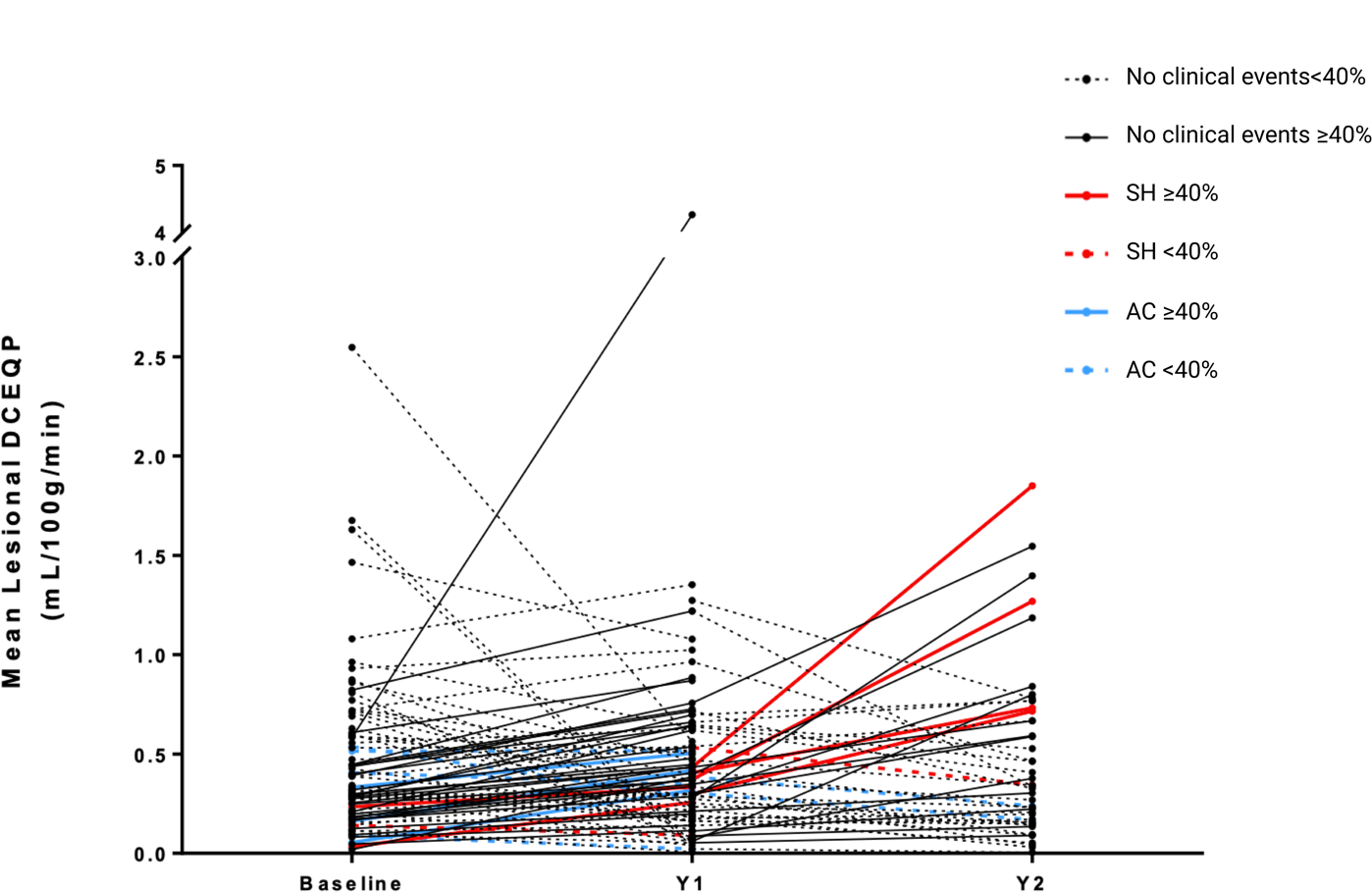
Mean lesional DCEQP at baseline, and at 1st and 2nd follow-up years. Black lines represent DCEQP pairs with no clinical events. Red lines represent DCEQP pairs with an SH event. Blue lines represent DCEQP pairs with AC events. DCEQP pairs meeting the threshold biomarker are solid lines. DCEQP pairs not meeting the biomarker threshold are dashed lines. Red lines represent yearly-epochs with SH; blue lines represent AC events, solid lines represent epoch-years that met the threshold biomarker of ΔDCEQP≥40%.

### Cases with AC and QSM yearly percent change < 6%

**Supplemental Figure 2A.**
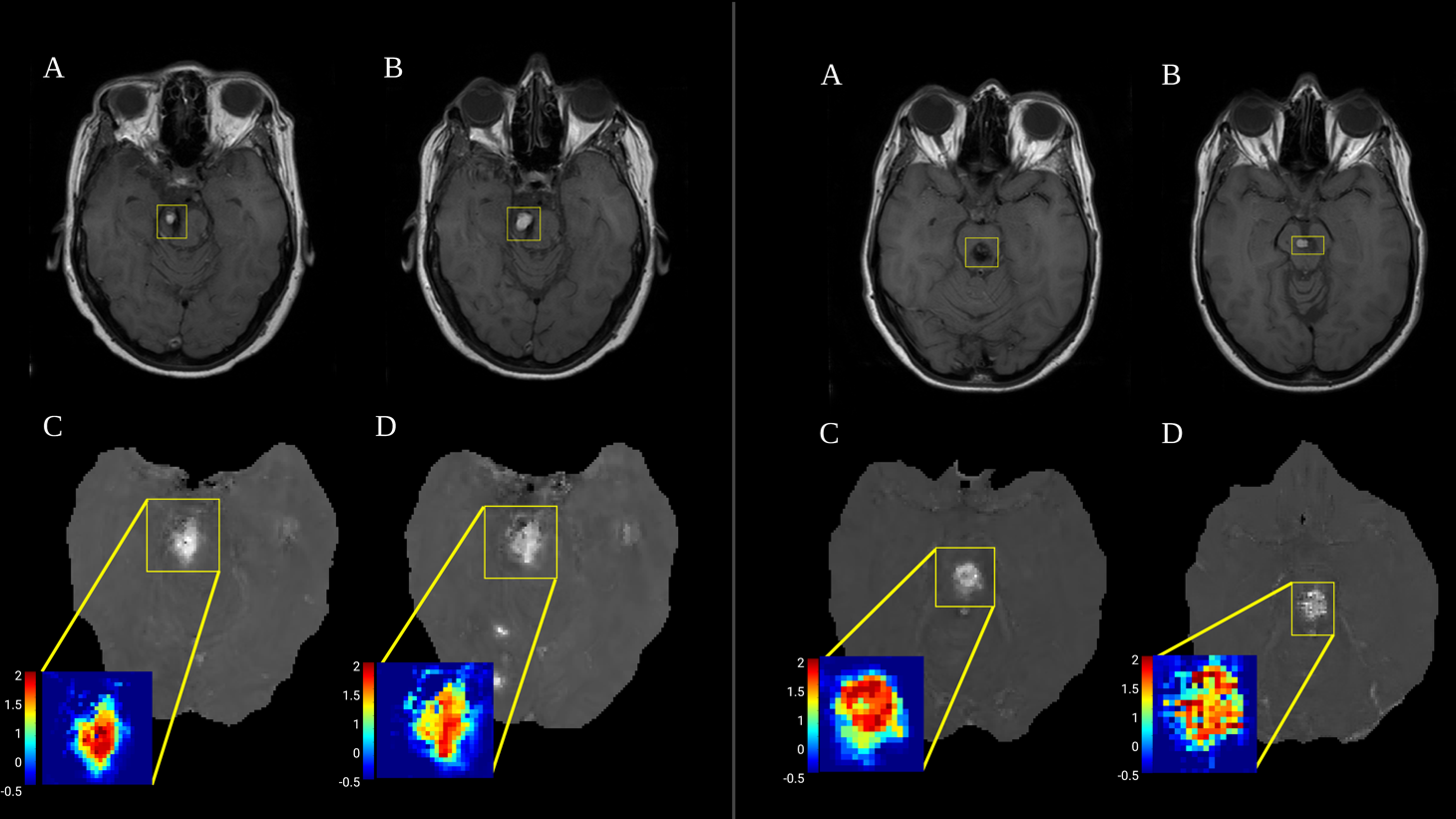
Change in prior bleed. For each case above: (A) Axial T1-weighted images showing the lesion of interest at the beginning of the epoch year; (B) Axial T1-weighted images showing the same lesion of interest at the end of the epoch year; (C) QSM acquisition on 1^st^ point in the epoch year with corresponding ROI selection for the lesion; (D) QSM acquisition on 2^nd^ point in the epoch year with corresponding lesion ROI selection.

The two cases above presented an asymptomatic change and exhibited an increase of QSM by < 6%. Neither case had new symptoms associated with the imaging change. In both instances, detailed review indicates that the increased blood signal in the lesion could reflect evolution of MRI appearance of previous hemorrhage rather than new bleeding.

**Supplemental Figure 2B.**
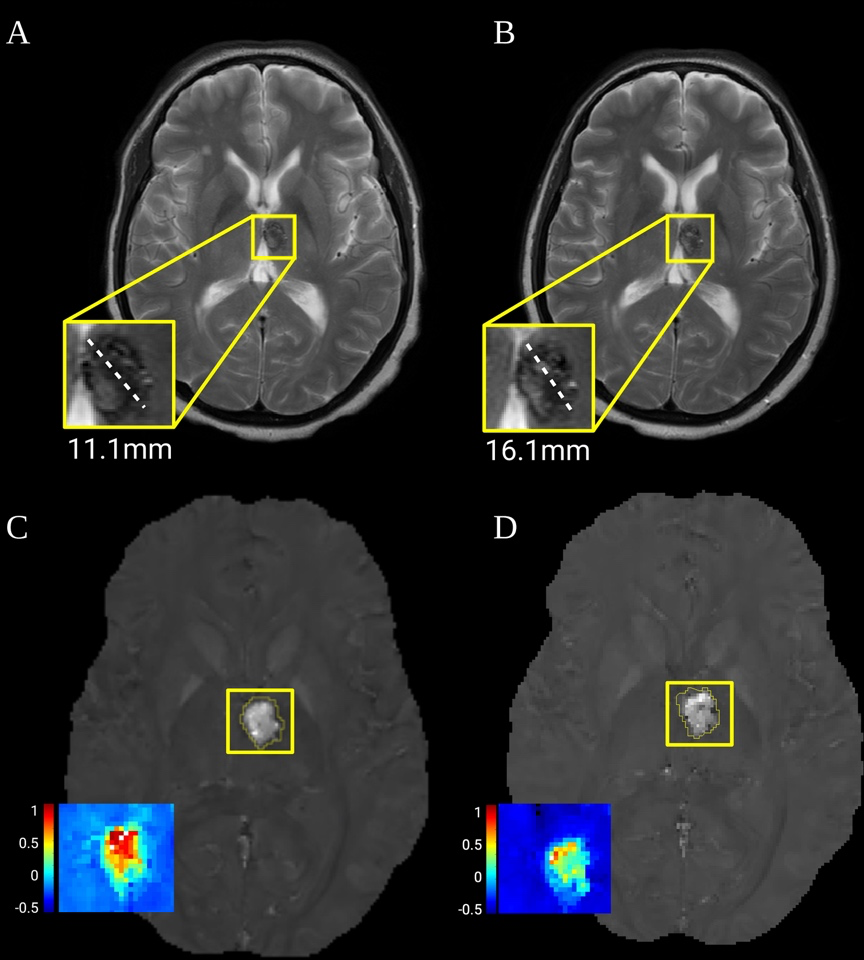
Asymptomatic lesion growth. (A) Axial T2-weighted images showing the lesion of interest with maximum axial diameter (11.1 mm) at the beginning of the epoch year; (B) Axial T2-weighted images showing the same lesion of interest size (16.11 mm) at the end of the epoch year; (C) QSM acquisition on 1^st^ point in the epoch year with corresponding ROI selection for the lesion; (D) QSM acquisition on 2^nd^ point in the epoch year with corresponding ROI selection for the lesion. There is no new bleeding, and the apparent increase in lesion diameter by >3mm triggering AC definition (documented by the site and verified by adjudication) may be attributable to differences in T2-weighted imaging techniques or diffusion of hemosiderin from the prior bleed.

### Timing of SH event within the follow-up epoch and percent change in mean lesional QSM and DCEQP among cases with SH who completed paired biomarker assessment

**Supplemental Figure 3.**
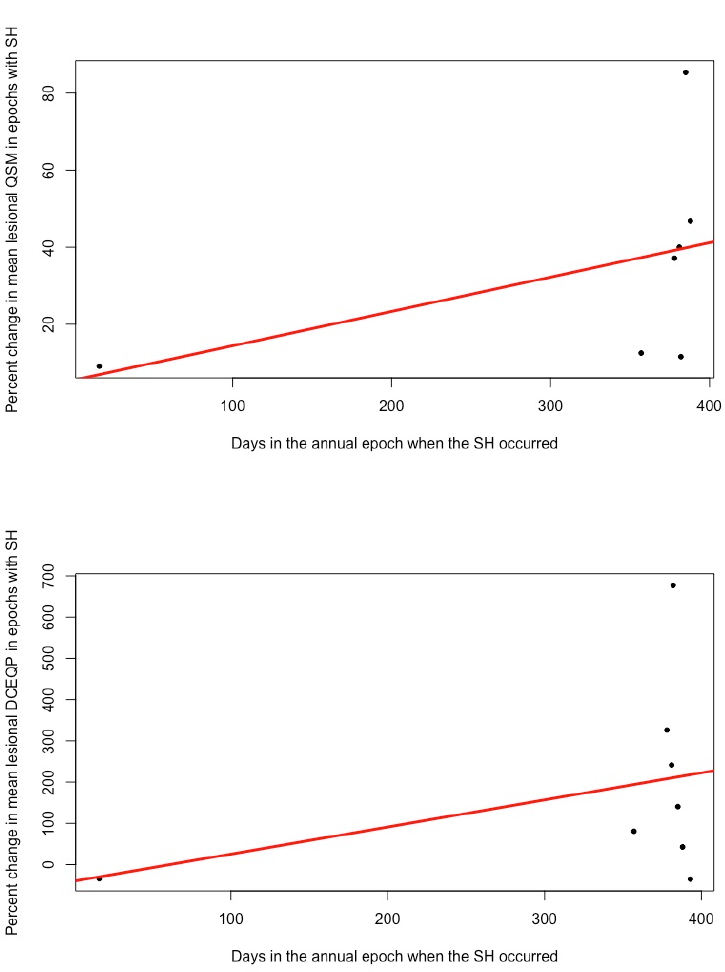
Percent change in QSM and DCEQP and timing of SH within the epoch, among cases with SH and paired biomarker assessments.

## Supplemental Tables

### Comparison of TR FUBV cases with and without paired biomarker assessments

**Supplemental Table 1:**
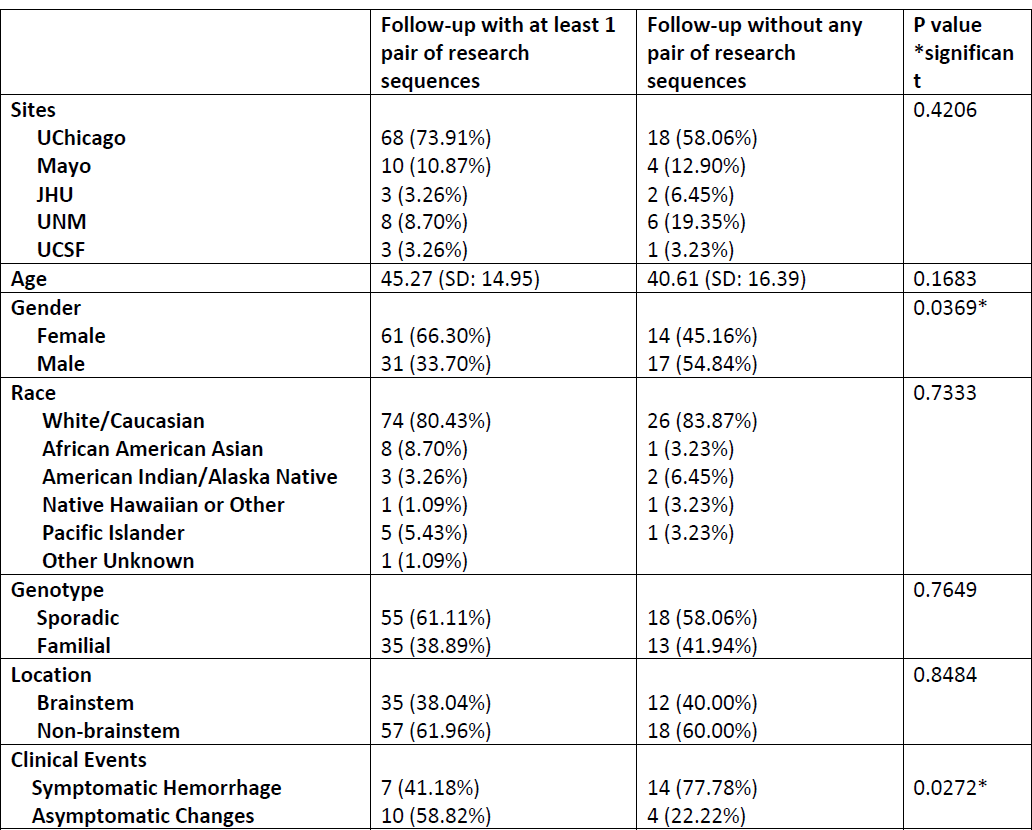
Features, demographics and clinical events for subjects contributing at least one pair of bio marker assessments compared to the ones without a pair of biomarker assessments. Age of subjects with and without MRI acquisition were compared using t-test. Chi-square test were used to compare proportions of other demographics distributions between groups. Significance was noted if P < 0.05. UChicago: University of Chicago, Mayo: Mayo Clinic Rochester, JHU: Johns Hopkins University, UNM: University of New Mexico, UCSF: University of California San Francisco.

|  | Follow-up with at least 1 pair of research sequences | Follow-up without any pair of research sequences | P value<br>*significant |
| --- | --- | --- | --- |
| **Sites** |  |  | 0.4206 |
| **UChicago** | 68 (73.91%) | 18 (58.06%) |  |
| **Mayo** | 10 (10.87%) | 4 (12.90%) |  |
| **JHU** | 3 (3.26%) | 2 (6.45%) |  |
| **UNM** | 8 (8.70%) | 6 (19.35%) |  |
| **UCSF** | 3 (3.26%) | 1 (3.23%) |  |
| **Age** | 45.27 (SD: 14.95) | 40.61 (SD: 16.39) | 0.1683 |
| **Gender** |  |  | 0.0369* |
| **Female** | 61 (66.30%) | 14 (45.16%) |  |
| **Male** | 31 (33.70%) | 17 (54.84%) |  |
| **Race** |  |  | 0.7333 |
| **White/Caucasian** | 74 (80.43%) | 26 (83.87%) |  |
| **African American** | 8 (8.70%) | 1 (3.23%) |  |
| **Asian** | 3 (3.26%) | 2 (6.45%) |  |
| **American Indian/Alaska Native** | 1 (1.09%) | 1 (3.23%) |  |
| **Native Hawaiian or Other Pacific Islander** | 5 (5.43%) | 1 (3.23%) |  |
| **Other Unknown** | 1 (1.09%) |  |  |
| **Genotype** |  |  | 0.7649 |
| **Sporadic** | 55 (61.11%) | 18 (58.06%) |  |
| **Familial** | 35 (38.89%) | 13 (41.94%) |  |
| **Location** |  |  | 0.8484 |
| **Brainstem** | 35 (38.04%) | 12 (40.00%) |  |
| **Non-brainstem** | 57 (61.96%) | 18 (60.00%) |  |
| **Clinical Events** |  |  | 0.0272* |
| **Symptomatic Hemorrhage** | 7 (41.18%) | 14 (77.78%) |  |
| **Asymptomatic Changes** | 10 (58.82%) | 4 (22.22%) |  |

**Supplemental Table 2:**
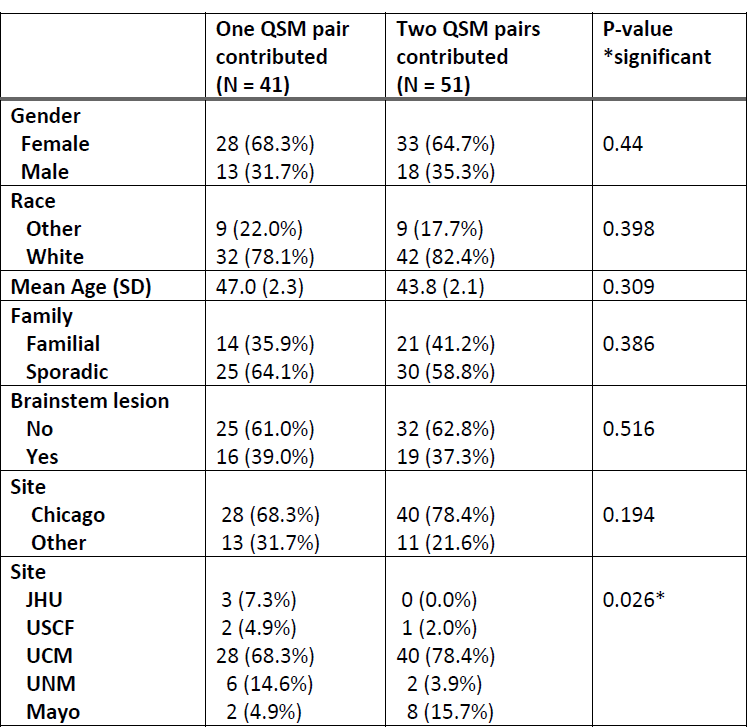
Features and demographics of subjects contributing to one and two QSM pairs and at multiple sites. Age of subjects with one and two QSM pairs were compared using t-test. Fisher’s Exact test was used to compare proportions of other demographics distributions between groups. Significance was noted if P < 0.05. UChicago: University of Chicago, Mayo: Mayo Clinic Rochester, JHU: Johns Hopkins University, UNM: University of New Mexico, UCSF: University of California San Francisco.

|  | One QSM pair<br>contributed<br>(N = 41) | Two QSM pairs<br>contributed<br>(N = 51) | P-value<br>*significant |
| --- | --- | --- | --- |
| **Gender** |  |  |  |
| Female | 28 (68.3%) | 33 (64.7%) | 0.44 |
| Male | 13 (31.7%) | 18 (35.3%) |  |
| **Race** |  |  |  |
| Other | 9 (22.0%) | 9 (17.7%) | 0.398 |
| White | 32 (78.1%) | 42 (82.4%) |  |
| **Mean Age (SD)** | 47.0 (2.3) | 43.8 (2.1) | 0.309 |
| **Family** |  |  |  |
| Familial | 14 (35.9%) | 21 (41.2%) | 0.386 |
| Sporadic | 25 (64.1%) | 30 (58.8%) |  |
| **Brainstem lesion** |  |  |  |
| No | 25 (61.0%) | 32 (62.8%) | 0.516 |
| Yes | 16 (39.0%) | 19 (37.3%) |  |
| **Site** |  |  |  |
| Chicago | 28 (68.3%) | 40 (78.4%) | 0.194 |
| Other | 13 (31.7%) | 11 (21.6%) |  |
| **Site** |  |  |  |
| JHU | 3 (7.3%) | 0 (0.0%) | 0.026* |
| USCF | 2 (4.9%) | 1 (2.0%) |  |
| UCM | 28 (68.3%) | 40 (78.4%) |  |
| UNM | 6 (14.6%) | 2 (3.9%) |  |
| Mayo | 2 (4.9%) | 8 (15.7%) |  |

**Supplemental Table 3:**
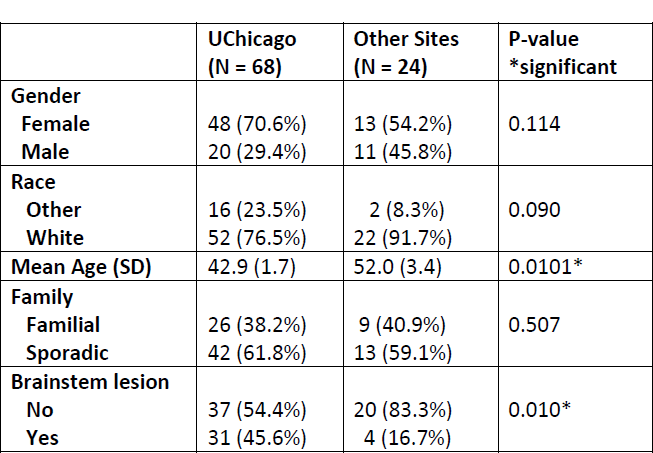
Features and demographics of subjects contributing to imaging biomarkers, enrolled at University of Chicago (largest enroller) and other sites. Age of subjects with biomarker imaging pairs enrolled at University of Chicago and other sites were compared using t-test. Fisher’s Exact test was used to compare proportions of other demographics distributions between groups. Significance was noted if P < 0.05.

### Trial Modeling

#### Sample size calculations for effect on percent change in mean lesional QSM or DCEQP during two years of follow-up

**Supplemental Table 4.** Analyses were conducted using time-averaged difference in % mean lesional QSM and DCEQP change between the two arms using a repeated measures analysis implemented as a linear mixed model during 2 years of follow-up (2 paired biomarker assessments), with 1:1 randomization. This is based on observed yearly-change standard deviations (SD) in two epochs, within-patient correlation changes scores between epoch 1 and epoch 2 (Rho), standard-deviation of the mean, power of 0.8 and alpha 0.05 for hypothesized interventional effect of 20%, 25% and 30%. Samples required to detect hypothesized effects are calculated per trial arm and for total trial population, not accounting for anticipated attrition. Based on QSM yearly percent change, detecting an effect of 20%, requires 92 (two-tailed) and 72 (one-tailed) subjects. For a 25% effect, 60 (two-tailed) and 46 (one-tailed) subjects are required, while 30% effect requires 42 (two-tailed) and 34 (one-tailed) subjects. Based on DCEQP yearly percent change, for 20%, 25% and 30% effect detection, 2062 (two-tailed) and 1624 (one tailed), 1322 (two-tailed) and 1040 (one-tailed), and 918 (two-tailed) and 724 (one-tailed) subjects are respectively required. The much larger sample size needed for DCEQP effect size is mostly due to the much greater variance in DCEQP measurements than QSM in the same subjects.

|  | Epoch 1 | Epoch 2 |  |  | 20% |  | 25% |  | 30% |  |
| --- | --- | --- | --- | --- | --- | --- | --- | --- | --- | --- |
|  | SD 1 | SD 2 | Rho | SD of mean | N per group |  | N per group |  | N per group |  |
|  |  |  |  |  | 2-sided | 1-sided | 1-sided | 2-sided | 2-sided | 1-sided |
| **QSM** | 40 | 72 | -0.4 | 33.5 | 46 | 36 | 23 | 60 | 21 | 17 |
| **DCEQP** | 281 | 208 | -0.15 | 162 | 1031 | 812 | 520 | 1322 | 459 | 362 |

#### Sample size calculations (patient-years per group) based on categorical outcome of annual SH rate and composite outcome (SH, AC or **Δ**QSM **≥**6%) rate

**Supplemental Table 5.**
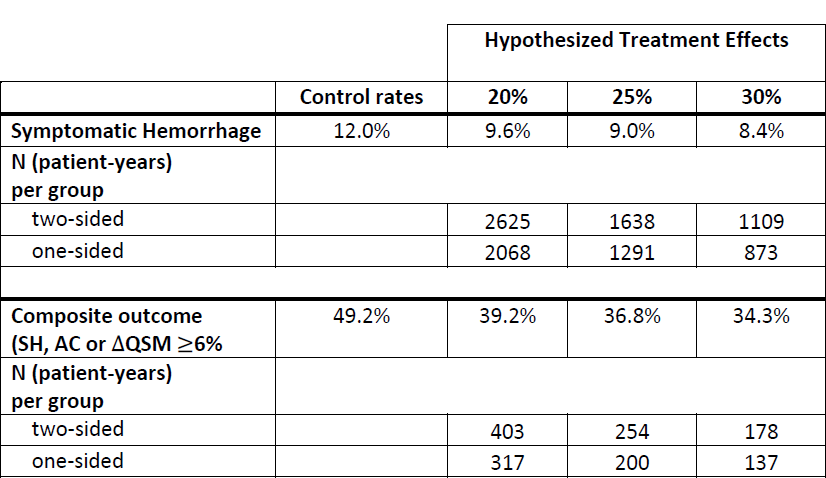
Assumptions are power 0.08 and alpha 0.05 for respective hypothesized therapeutic effects of 20%, 25% and 30% change in the annual frequency of SH (12.0%) and composite categorical outcome (49.2%) with 1:1 randomization. A 20% reduction in SH rate requires 5250 (two-tailed) or 4136 (one-tailed) patient-years; a 25% reduction in SH rate requires 3276 (two-tailed) or 2582 (one-tailed) patient-years and a reduction of 30% in SH rates requires 2218 (two-tailed) or 1746 (one-tailed) patient-years. Reduction of 20% in composite outcome rate could be detected in 806 (two-tailed) or 634 (one-tailed) patient-years; reduction of 25% is detected in 508 (two-tailed) or 400 (one-tailed) patient-years; and a reduction of 30% is detected with in 356 (two-tailed) or 274 (one-tailed) patient-years. With this modeling, half as many subjects are needed if each subject is followed for two years, assuming the same frequency of events in both years.

|  |  | **Hypothesized Treatment Effects** |  |  |
| --- | --- | --- | --- | --- |
|  | **Control rates** | **20%** | **25%** | **30%** |
| **Symptomatic Hemorrhage** | 12.0% | 9.6% | 9.0% | 8.4% |
| **N (patient-years)**<br>**per group** |  |  |  |  |
| two-sided |  | 2625 | 1638 | 1109 |
| one-sided |  | 2068 | 1291 | 873 |
| **Composite outcome**<br>**(SH, AC or ΔQSM ≥6%)** | 49.2% | 39.2% | 36.8% | 34.3% |
| **N (patient-years)**<br>**per group** |  |  |  |  |
| two-sided |  | 403 | 254 | 178 |
| one-sided |  | 317 | 200 | 137 |

